# Outcomes of home-based versus facility-based care for mild diphtheria during a large epidemic in Kano State, Nigeria: a retrospective matched cohort study

**DOI:** 10.64898/2026.04.10.26350586

**Authors:** Jonathan Polonsky, Suleiman Hudu, Khadidja Uthman, Yves Katuala, Philip Eghosa Evbuomwan, Hashim Juma Omer Osman, Abdulwahab Kabir Sulaiman, Ichola Ismael Adjaho, Cheick Oumar Doumbia, Etienne Gignoux, Franck Ale

**Affiliations:** Epicentre, Paris, France; Geneva Centre of Humanitarian Studies, Faculty of Medicine, University of Geneva, Geneva, Switzerland; Médecins Sans Frontières, Operational Centre West and Central Africa (MSF WaCA); Kano State Ministry of Health, Kano State, Nigeria

**Keywords:** diphtheria, decentralised care, home-based care, outbreak response, Nigeria, cohort study

## Abstract

**Background:** During Nigeria’s largest recorded diphtheria outbreak, hospital capacity in Kano State was rapidly overwhelmed. Médecins Sans Frontières introduced home-based care (HBC) for patients with mild disease to prioritise facility-based care for severe cases. We assessed whether HBC was non-inferior to facility-based treatment in terms of mortality, sequelae, and household transmission.

**Methods:** We conducted a retrospective matched cohort study. Mild diphtheria cases treated between January 2023 and May 2024 were matched 1:1 by treatment modality (HBC or diphtheria treatment centre [DTC]) on sex, age group, vaccination status, and residence. Conditional logistic regression estimated the association between treatment modality and mortality, with robustness assessed through propensity score weighting, sensitivity analyses, and E-value computation.

**Findings:** Of 990 sampled patients, 678 (367 HBC, 311 DTC) were enrolled (68·5%). After adjustment, treatment modality was not independently associated with mortality (HBC vs. DTC: aOR 0·40, 95% CI 0·13–1·30), with similar estimates across sensitivity analyses (E-value 4·40). Clinical complications were the strongest predictor of death (aOR 23·1, 95% CI 1·73–307). Vaccination was protective (aOR 0·28, 95% CI 0·08– 0·94) and treatment delay of four or more days increased mortality (aOR 4·15, 95% CI 1·23–14·0). HBC was not associated with increased household transmission or long-term sequelae.

**Interpretation:** Vaccination and early treatment, rather than care setting, were the main determinants of survival. When supported by clinical triage and structured follow-up, decentralised care can be used to manage mild cases during diphtheria epidemics in settings with constrained hospital capacity.

**Funding:** Médecins Sans Frontières, West and Central Africa.

**Research in context:** *Evidence before this study:* We searched PubMed and Google Scholar for articles published between January 1, 2000, and February 28, 2026, using combinations of the terms “diphtheria”, “outbreak”, “home-based care”, “outpatient”, “ambulatory”, “community care”, and “decentralised care”. We found no published studies evaluating any form of decentralised or home-based clinical management for diphtheria. The existing literature on diphtheria case management is confined to facility-based settings: outbreak reports from multiple affected countries describe hospital-based treatment with diphtheria antitoxin (DAT) and antibiotics, and a systematic review pooled epidemiological and clinical data from historical outbreaks. Decentralised care models have been evaluated for other epidemic-prone diseases, including a measles epidemic in the Democratic Republic of the Congo (DRC) where decentralised management reduced mortality among children, and Ebola virus disease outbreaks in DRC where decentralised treatment centres were piloted to improve geographic access, though with limited outcome data. No study has assessed whether patients with diphtheria can be safely managed outside hospital settings.

*Added value of this study:* No prior evaluation of home-based care for diphtheria has been published. Using a retrospective matched cohort design with 678 patients during the largest diphtheria outbreak in Africa in decades, we found no evidence that home-based care increased mortality, long-term complications, or household transmission compared with facility-based care, and acceptability was high among patients in both groups. The study also provides one of the largest datasets on household transmission of diphtheria in an urban epidemic setting, finding no evidence that home-based care increased secondary transmission, and showing that vaccination status of the index case was the main factor influencing spread within the household.

*Implications of all the available evidence:* Provided that triage is reliable, antibiotics are started promptly, and a functioning referral pathway exists, mild diphtheria can be managed safely at home during large epidemics. This approach preserves limited hospital and DAT resources for patients with moderate or severe disease, shortens treatment delays, and is acceptable to patients. Given ongoing outbreaks across West and Central Africa and persistent DAT supply constraints, decentralised care warrants inclusion in epidemic preparedness.

## Introduction

Conventional management of diphtheria requires hospitalisation for administration of diphtheria antitoxin (DAT), antibiotic therapy, and monitoring for toxin-mediated complications such as myocarditis and polyneuropathy.^1,2^ This assumes the availability of inpatient capacity, trained staff, and a reliable DAT supply, conditions rarely met during large outbreaks in resource-constrained settings. In Bangladesh, nine dedicated diphtheria treatment centres (DTCs) struggled to meet demand during the 2017–19 Rohingya refugee outbreak despite substantial international support.^3^ In Yemen, DAT shortages forced clinicians to ration treatment.^4^ When the current epidemic struck Kano State, Nigeria, in late 2022, hospital capacity was overwhelmed within months.^5,6^

Nigeria’s ongoing diphtheria epidemic is the largest in Africa in decades, with over 61,000 suspected cases nationally and approximately 25,000 confirmed cases in Kano State alone.^5,7^ A companion population-based survey estimated that community attack rates in the most affected areas were approximately four times higher than facility-based surveillance suggested, with the majority of deaths occurring at home.^8^ Facility-based data from Kano documented a case fatality rate of 4·5% among 18,320 surveillance-captured cases.^5^

In response to rapidly increasing cases during mid-2023, Médecins Sans Frontières (MSF), in collaboration with the Kano State Ministry of Health, introduced home-based care (HBC) for patients assessed as having mild diphtheria without signs of complications. These patients received oral antibiotics and were discharged home with follow-up visits on days 3 and 7, weekly outpatient reassessment, and a clear referral pathway to a DTC if they deteriorated. Patients with signs of severe disease (respiratory distress, extensive pharyngeal membrane, or suspected toxin-mediated complications) were admitted to DTCs for inpatient management including DAT. Over the 15-month response period, MSF managed 14,707 patients, of whom 6,696 (45·5%) received HBC.

Severity and complication risk in diphtheria are determined largely by the extent and duration of toxin exposure. ^1,2^ Patients with mild pharyngeal disease and limited pseudomembrane who receive early antibiotics to halt toxin production might plausibly be managed safely outside hospital. However, the risk of clinical deterioration (particularly late-onset myocarditis, which can occur days to weeks after initial presentation) and the potential for increased household transmission represent key safety concerns.^2,9^

We conducted a retrospective matched cohort study to evaluate the safety and effectiveness of HBC compared with DTC-based care for mild diphtheria during the Kano State epidemic. We assessed mortality, long-term sequelae, household transmission, and patient-reported acceptability, and examined clinical and care-delivery factors associated with these outcomes.

## Methods

### Study design and setting

This was a retrospective matched cohort study conducted in four of the most affected LGAs of Kano State (Dawakin Tofa, Fagge, Nassarawa, and Ungogo) between September and October 2024.

### Participants and matching

We identified all patients with a clinical diagnosis of mild diphtheria recorded in MSF registries who had sought treatment between 1 January 2023 and 31 May 2024. From these records, 496 patients managed as inpatients in DTCs and classified as mild at initial assessment were selected by simple random sampling. An equal number of HBC patients were then randomly selected using 1:1 matching on sex, age group (within five-year bands), vaccination status, and LGA of residence. Patients who could not be traced after two contact attempts or who declined participation were excluded without replacement.

Matching was designed to ensure baseline comparability on the main demographic and clinical characteristics that might confound the association between treatment setting and outcomes. Treatment modality was determined by clinical triage at presentation: patients without respiratory distress and with limited pseudomembrane extension were eligible for HBC, while those with any signs suggesting moderate or severe disease were admitted to DTCs.

### Data collection

Data were collected through structured household interviews using KoboCollect^10^ on encrypted tablets, supplemented by review of MSF clinical records. The questionnaire captured demographics, vaccination status, symptom onset and treatment timelines, clinical outcomes, long-term sequelae, and diphtheria status for all household members. A dedicated section assessed patient-reported acceptability using a five-point Likert scale across three dimensions: perceived improvement, acceptability of the care model, and perceived quality. Clinical records were linked to interview data to verify treatment history, clinical presentation, and complication status.

### Definitions

A diphtheria case was any individual recorded in MSF databases with a clinical diagnosis of suspected diphtheria, regardless of laboratory confirmation. Mild disease was defined by the absence of respiratory distress and limited pseudomembrane extension on clinical examination. Vaccination referred to receipt of at least one dose of a diphtheria-containing vaccine according to card-verified dates (and caregiver recall in absence of cards).

Secondary household cases were defined as household members who developed diphtheria-compatible symptoms within 2–14 days after the index case, with a broader 2–30 day window examined in sensitivity analysis. Clinical complications comprised acute manifestations consistent with toxin-mediated disease, including dyspnoea requiring referral, suspected myocarditis, neurological involvement, or other clinician-documented serious events.

### Outcomes

Primary outcomes were all-cause mortality, long-term sequelae (assessed at the time of interview), intra-household secondary attack rate (SAR), and patient-reported acceptability.

### Sample size

We estimated that 992 patients (496 per group) would provide 80% power to detect a two-fold difference in mortality between treatment groups, assuming a baseline CFR of 2·5% in the DTC group, a two-sided alpha of 0·05, and 10% non-response.

### Statistical analysis

All analyses were conducted in R (version 4.3). Descriptive statistics summarised the study population. Proportions are presented with 95% confidence intervals (CIs) using binomial exact methods for SARs and design-adjusted methods for mortality.

As the study used a matched design, individuals were grouped into matched subclasses. Conditional logistic regression, stratified by subclass, was used to estimate odds ratios (ORs) for mortality and other binary outcomes. *A priori* covariates included treatment modality, age, vaccination status, treatment delay, and clinical complications. Treatment delay was modelled both as a continuous variable and dichotomised at ≥4 days, based on an observed non-linear increase in mortality beyond this threshold.

Calendar time was assessed as a potential confounder, given that HBC was introduced partway through the outbreak and care-seeking patterns and system capacity may have evolved over time. It was parameterised as a continuous variable (onset date), using restricted cubic splines, and as categorical epidemic phases, and retained in the model only if it materially altered the treatment modality estimate. We also tested whether the treatment modality effect varied by epidemic phase using an interaction term.

Household transmission was analysed using household member data reported during interviews pertaining to the index cases, with SAR calculated as secondary cases divided by total household members at risk. Multivariable logistic regression with cluster-robust standard errors was used to examine risk factors, including treatment modality of the index case, age, sex, vaccination status, household size, treatment delay, and complications.

Acceptability was analysed using linear mixed-effects models with matched subclass as a random intercept, reported for each Likert dimension and as a composite mean score. All inferential analyses used complete cases.

### Sensitivity and robustness analyses

We conducted three sensitivity analyses. First, we excluded the eight DTC patients who developed clinical complications and reran the conditional logistic regression. Second, we estimated the treatment effect using stabilised inverse probability of treatment weighting (IPTW), trimmed at the 1st and 99th percentiles. Weights were derived from a model including age, sex, vaccination status, LGA of residence, and household size as predictors of treatment allocation. Third, we computed E-values for the treatment modality, vaccination, and treatment delay estimates to quantify the minimum unmeasured confounding required to explain each observed association.^11^

### Ethics

Approval was granted by the MSF Ethics Review Board and the Kano State Ministry of Health Ethics Committee. Informed consent was obtained from all participants or their guardians. Data were anonymised and stored in accordance with data protection regulations.

## Results

### Participants

From 3,462 eligible patients in MSF registries, 990 were randomly selected (496 DTC, 494 HBC). Of these, 678 (68·5%) were traced and enrolled: 367 HBC and 311 DTC (Table 1, Figure 1). The temporal distribution of the two groups differed, reflecting the introduction of HBC partway through the outbreak: median symptom onset was 3 July 2023 for DTC patients and 4 September 2023 for HBC patients (Figure 2).

**Table 1.** Characteristics of enrolled participants by treatment modality, Kano State diphtheria outbreak, Nigeria, 2023–24.

| Characteristic | HBC<br>(n=367) | DTC<br>(n=311) | Total<br>(n=678) |
| --- | --- | --- | --- |
| **LGA** |  |  |  |
| Dawakin Tofa | 20 (5%) | 12 (4%) | 32 (5%) |
| Fagge | 108 (29%) | 86 (28%) | 194 (29%) |
| Nassarawa | 46 (13%) | 41 (13%) | 87 (13%) |
| Ungogo | 193 (53%) | 172 (55%) | 365 (54%) |
| **Sex** |  |  |  |
| Female | 229 (62%) | 194 (62%) | 423 (62%) |
| Male | 138 (38%) | 117 (38%) | 255 (38%) |
| **Age group** |  |  |  |
| 0–5 years | 38 (10%) | 36 (12%) | 74 (11%) |
| 6–10 years | 140 (38%) | 111 (36%) | 251 (37%) |
| 11–15 years | 99 (27%) | 80 (26%) | 179 (26%) |
| ≥16 years | 90 (25%) | 84 (27%) | 174 (26%) |
| **Vaccinated** |  |  |  |
| No | 131 (36%) | 95 (31%) | 226 (33%) |
| Yes | 236 (64%) | 216 (69%) | 452 (67%) |
| **Comorbidity** | 0 | 6 (2%) | 6 (1%) |
| **Complication** | 0 | 8 (3%) | 8 (1%) |
| **Median household size (IQR)** | 7 (5–10) | 8 (5–12) | 7 (5–10) |
| **Median treatment delay, days (IQR)** | 2 (1–3) | 2 (1–3) | 2 (1–3) |
| **Outcome** |  |  |  |
| Recovered | 353 (96%) | 286 (92%) | 639 (94%) |
| Died | 7 (2%) | 17 (5%) | 24 (4%) |
| Still unwell | 6 (2%) | 8 (3%) | 14 (2%) |

*HBC = home-based care. DTC = diphtheria treatment centre. LGA = local government area. IQR = interquartile range.*

**Figure 1.**
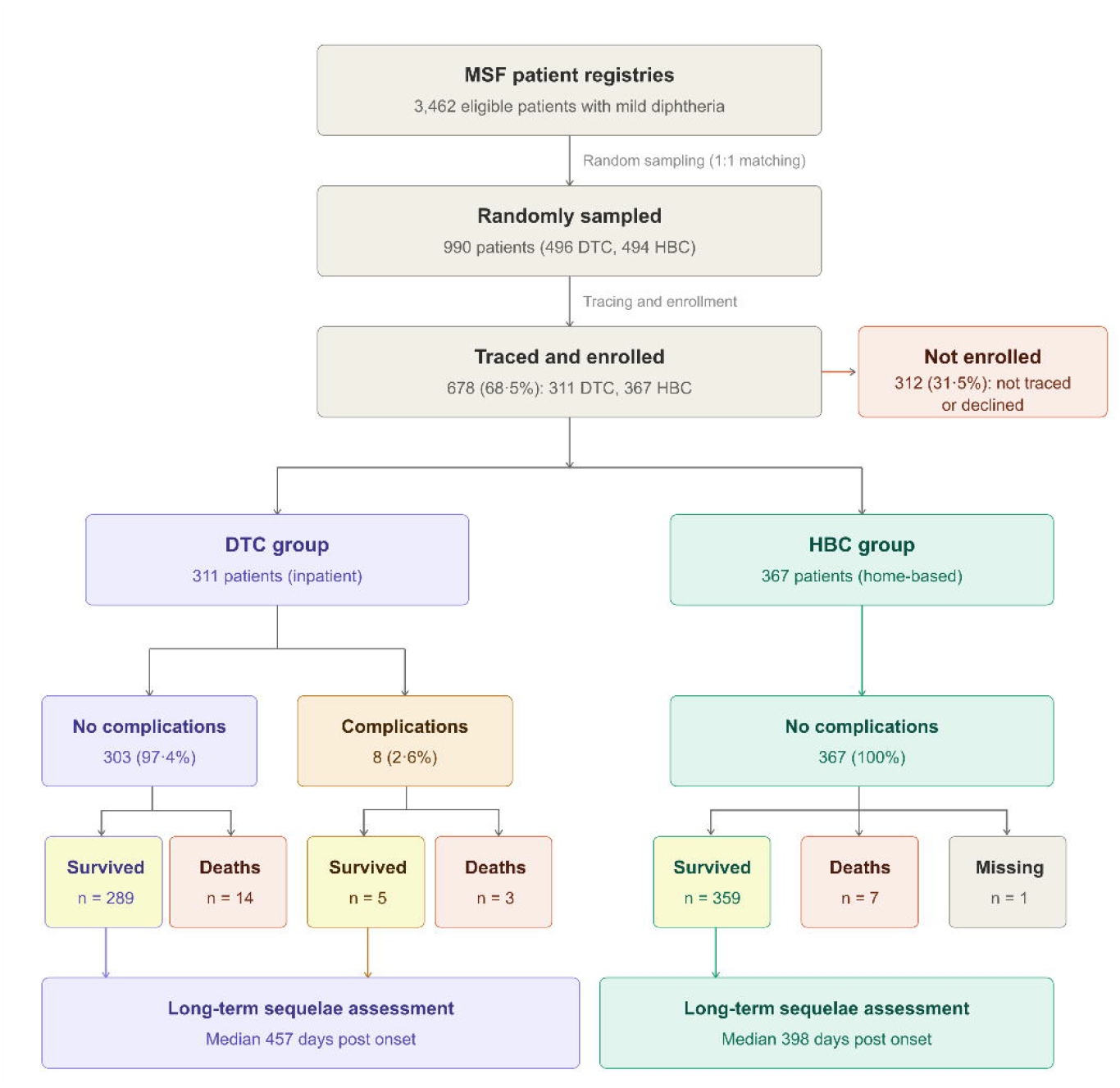
Flow of participants from the MSF case registry through eligibility screening, household tracing, enrolment, and analysis, by treatment modality, Kano State diphtheria outbreak, Nigeria, 2023–24.

**Figure 2.**
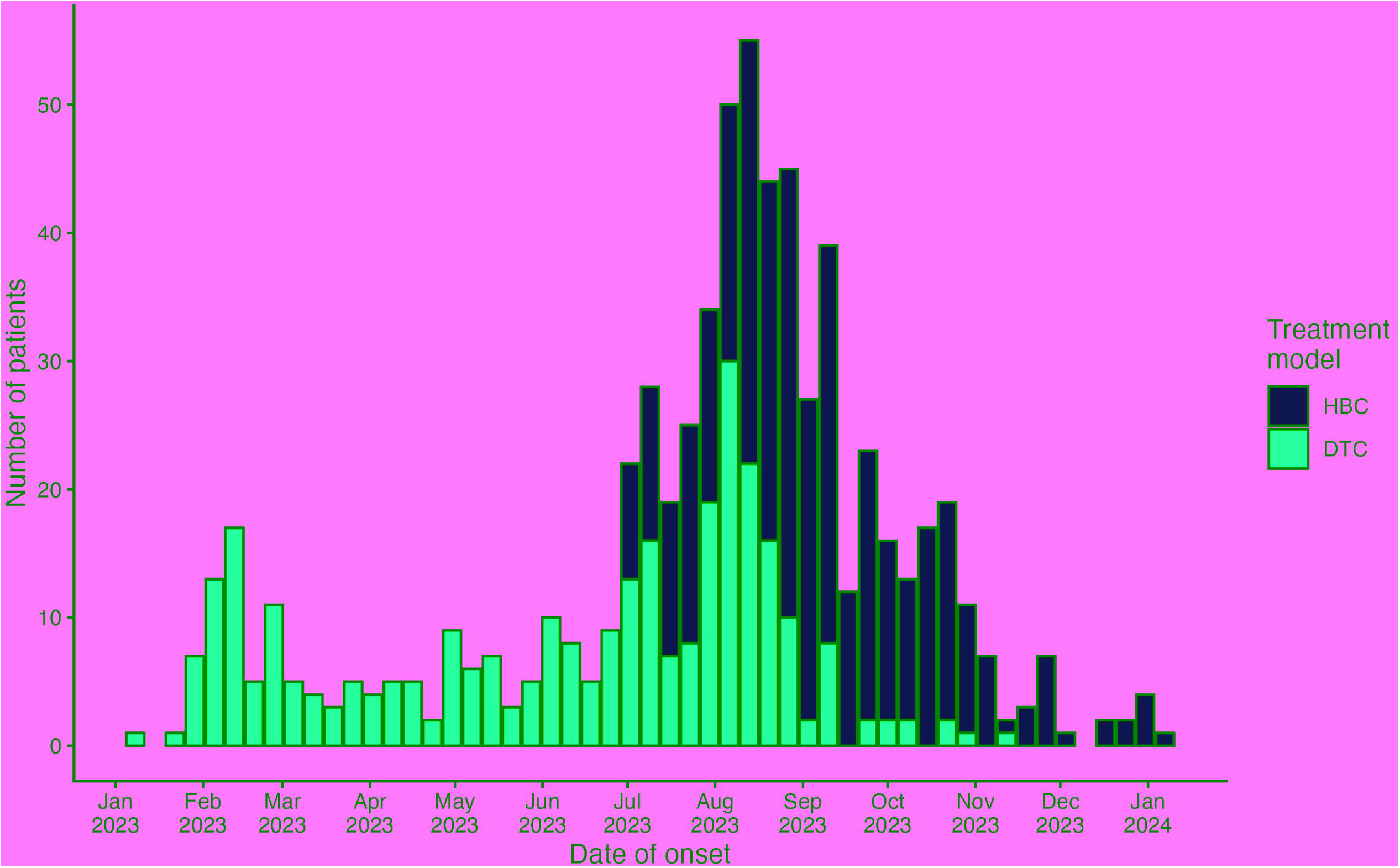
Epidemic curve of enrolled participants by week of symptom onset and treatment modality, Kano State diphtheria outbreak, Nigeria, 2023–24. *Each bar represents one epidemiological week. HBC = home-based care. DTC = diphtheria treatment centre*.

The median age was 11 years (IQR 8–16) and 62·4% were female. Reported vaccination coverage was 66·7% (95% CI 63·3–70·1), similar across treatment groups (64·3% HBC vs 69·5% DTC). No comorbidities were documented among HBC patients, whereas 1·9% of DTC patients had comorbidities (principally peptic ulcer disease). Similarly, no clinical complications were recorded among HBC patients, whereas 2·6% of DTC patients developed complications (including acute kidney injury, thrombocytopenia, airway obstruction, and heart failure). Median household size was slightly larger among DTC patients (8 vs 7, p=0·011). Median treatment delay was 2 days (IQR 1–3) in both groups (p=0·46).

### Mortality

There were 24 deaths among 678 patients (CFR 3·5%, 95% CI 2·3–5·2): 7 among 367 HBC patients (1·9%, 95% CI 0·5–3·3) and 17 among 311 DTC patients (5·5%, 95% CI 3·0–8·0) (Table 1). In unadjusted conditional logistic regression, DTC treatment was associated with higher odds of death (HBC vs. DTC: aOR 0·32, 95% CI 0·12–0·86).

After adjustment, treatment modality was not independently associated with mortality (HBC vs. DTC: aOR 0·40, 95% CI 0·13–1·30, p=0·128), with results consistent across analytical approaches supporting non-inferiority of HBC (Table 2). Clinical complications were the strongest independent predictor of death (aOR 23·1, 95% CI 1·73–307). Vaccination was protective (aOR 0·28, 95% CI 0·08–0·94), and increasing age was associated with lower mortality (aOR 0·73 per year, 95% CI 0·55–0·96).

**Table 2.** Conditional logistic regression of predictors of mortality among enrolled participants, Kano State diphtheria outbreak, Nigeria, 2023–24.

| Predictor | Unadjusted OR (95% CI) | Adjusted OR (95% CI) | p |
| --- | --- | --- | --- |
| **Treatment modality** |  |  |  |
| HBC | - | - | 0.128 |
| DTC | 0.32 (0.12–0.86) | 0.40 (0.13–1.30) |  |
| **Age** |  |  |  |
| Per year increase | 0.75 (0.59–0.95) | 0.73 (0.55–0.96) | 0.024* |
| **Vaccination status** |  |  |  |
| Unvaccinated | - | - | 0.040* |
| Vaccinated | 0.25 (0.09–0.72) | 0.28 (0.08–0.94) |  |
| **Complication** |  |  |  |
| Absent | - | - | 0.018* |
| Present | 15.68 (2.22–110.9) | 23.05 (1.73–307) |  |
| **Treatment delay** |  |  |  |
| <4 days | - | - | 0.022* |
| ≥4 days | 3.55 (1.30–9.70) | 4.15 (1.23–14.0) |  |

*HBC = home-based care. DTC = diphtheria treatment centre. OR = odds ratio. CI = confidence interval.*

Treatment delay showed a non-linear association with mortality. Each additional day increased the adjusted odds of death by 16% (aOR 1·16 per day, 95% CI 1·05–1·29), and delays of four or more days were associated with a four-fold increase in mortality (aOR 4·15, 95% CI 1·23–14·0).

Calendar time was not associated with mortality and did not materially alter the treatment modality estimate; it was therefore excluded from the final model. There was no evidence that the treatment modality effect varied by epidemic phase (interaction p>0·1).

### Robustness of the treatment modality estimate

The finding of no independent association between treatment modality and mortality was consistent across analytical approaches. Excluding the eight DTC patients with clinical complications did not materially change the estimate (HBC vs DTC: aOR 0·40, 95% CI 0·13–1·30, p=0·127). The IPTW analysis gave a marginally stronger estimate (HBC vs DTC: aOR 0·33, 95% CI 0·13–0·81), which reached statistical significance. The E-value for the treatment modality estimate (HBC vs DTC: aOR 0·40) was 4·40 for the point estimate, with a lower bound of 1·00. E-values for the vaccination effect (6·60) and treatment delay effect (7·77) indicate substantially greater robustness to unmeasured confounding.

### Long-term sequelae

Among 653 surviving patients assessed between 251 and 624 days after disease onset (median 421 days), 14 (2·1%) reported long-term sequelae: five had experienced relapse or reinfection, two each continued to experience neuritis and otitis media, and one patient each reported persistent respiratory difficulty and malnutrition. Treatment modality was not associated with sequelae (HBC vs DTC: aOR 0·80, 95% CI 0·26– 2·50), but acute complications were strongly predictive (aOR 30·5, 95% CI 4·36–214).

### Household transmission

Using the primary 2–14 day window, 137 secondary cases were identified among 5,066 household members of the 678 index cases (SAR 2·7%, 95% CI 2·28–3·19). SARs were 2·2% among household members of HBC patients and 3·2% among those of DTC patients (Table 3). There was no evidence that HBC increased household transmission (HBC vs DTC: aOR 0·81, 95% CI 0·60–1·10).

**Table 3.** Household secondary attack rates by treatment modality and time window from index case symptom onset, Kano State diphtheria outbreak, Nigeria, 2023–24.

| Time window | HBC |  | DTC |  | Overall |  |
| --- | --- | --- | --- | --- | --- | --- |
|  | Cases/N | SAR (95% CI) | Cases/N | SAR (95% CI) | Cases/N | SAR (95% CI) |
| 2–14 days | 58/2,608 | 2.2%<br>(1.7–2.9) | 79/2,458 | 3.2%<br>(2.6–4.0) | 137/5,066 | 2.7%<br>(2.3–3.2) |
| 2–30 days | 77/2,608 | 3.0%<br>(2.4–3.7) | 91/2,458 | 3.7%<br>(3.0–4.5) | 168/5,066 | 3.3%<br>(2.8–3.9) |
| Any timing | 176/2,608 | 6.8%<br>(5.8–7.8) | 212/2,458 | 8.6%<br>(7.5–9.8) | 388/5,066 | 7.7%<br>(6.9–8.4) |

*HBC = home-based care. DTC = diphtheria treatment centre. SAR = secondary attack rate, calculated as the number of household members developing symptoms within the specified window divided by the total number of susceptible household contacts. Index cases are excluded from both numerator and denominator. CIs calculated using exact binomial methods.*

In sensitivity analyses, 168 secondary cases were identified using the 2–30 day window (SAR 3·3%, 95% CI 2·84–3·85) and 388 using an unrestricted window (SAR 7·7%, 95% CI 6·94–8·43). The point estimate indicated lower transmission in HBC households across all three windows, though none reached statistical significance.

### Acceptability

Treatment was highly acceptable in both groups. Mean scores were 4·8 out of 5 for DTC care and 4·7 out of 5 for HBC across all three Likert dimensions, and acceptability scores were comparable between groups, with a small difference favouring DTC (ratio 1·12, 95% CI 1·03–1·22). Earlier treatment was associated with higher acceptability (ratio 1·14, 95% CI 1·02–1·27), and experiencing complications with lower acceptability (ratio 0·68, 95% CI 0·46–1·00).

## Discussion

During a large diphtheria epidemic that overwhelmed hospital capacity in Kano State, home-based care for patients with mild disease produced outcomes comparable to facility-based treatment. We observed no increase in mortality, sequelae, household transmission, or acceptability concerns associated with HBC. Survival was most strongly associated with treatment delay, vaccination status, and complications, with no independent association observed for care setting.

### Decentralised care for diphtheria

Textbook management of diphtheria centres on hospitalisation, DAT administration, and close monitoring for cardiac and neurological complications.^1,2^ This guidance was developed when most cases were managed in well-resourced hospital settings in high-income countries, and it remains the standard of care for moderate and severe disease. During large outbreaks in settings with limited health infrastructure, however, strict adherence to facility-based management creates a bottleneck that can paradoxically worsen outcomes by delaying treatment for some patients. Decentralised approaches have reduced mortality in measles outbreaks in the DRC primarily by shortening treatment delays,^12^ and have improved geographic access to care during Ebola and cholera responses.^13–15^ For diphtheria specifically, no evaluation of outpatient or community-based care has previously been published.

The toxin-mediated pathophysiology of diphtheria makes it particularly well suited for decentralised care. The risk of serious complications is largely determined by the quantity and duration of toxin exposure before treatment begins.^1,2,9^ Early antibiotics halt toxin production, and while DAT neutralises circulating free toxin, it cannot reverse damage already done.^2^ For patients with mild pharyngeal disease and limited pseudomembrane who present promptly, the risk of late complications is low, making community-based follow-up a clinically plausible approach. No HBC patient developed clinical complications, and the CFR among HBC patients (1·9%) was lower than the 4·5% recorded in Kano facility surveillance, although this included cases across the severity spectrum.^5^

### Mortality

In the adjusted analysis, treatment timing and vaccination status, rather than care setting, were the principal determinants of survival. After accounting for matching and key covariates, treatment modality was not associated with mortality (aOR 0·40, 95% CI 0·13–1·30). The higher crude CFR among DTC patients likely reflected residual confounding by severity. All clinical complications and comorbidities occurred in DTC patients, suggesting that triage did not fully exclude patients with some degree of undetected severity at presentation. The IPTW estimate (0·33, 95% CI 0·13–0·81) is consistent with this interpretation, though propensity score methods cannot adjust for unmeasured severity differences such as pseudomembrane extent.

Treatment delay was a strong and consistent predictor of death. Each additional day increased the odds of mortality by 16%,with delays of four or more days associated with a four-fold increase in mortality. This finding corroborates the companion survey, which estimated a 33-fold increase in mortality risk with delays beyond three days,^8^ and is consistent with established toxin kinetics and historical data showing DAT is most effective within 48 hours.^3,9,16^ Decentralised care can directly improve treatment timing by bringing the point of care closer to the patient.

Vaccination was another strong protective factor (aOR 0·28; E-value 6·60), consistent with the pooled estimate from a systematic review that three doses of diphtheria toxoid provide 87% protection against symptomatic disease.^9^ Closing vaccination gaps remains the most effective long-term strategy for preventing diphtheria mortality.

### Household transmission

A legitimate concern with managing an infectious respiratory disease at home is the potential for increased household transmission. The SAR was 2·7% overall, and was lower among household members of HBC patients (2·2%) than DTC patients (3·2%), a pattern consistent across all three time windows and robust to adjustment. This may reflect rapid reduction in infectiousness following antibiotic treatment, with the MSF protocol including antibiotic prophylaxis as well as household-level education and shielding guidance.

### Acceptability

Patient-reported acceptability was high for both care models. The clinically trivial 0·1-point advantage for DTC care suggests patients valued both approaches, with a slight preference for the perceived security of a hospital environment. Earlier treatment was associated with higher acceptability scores, consistent with the importance patients place on timely access to care.

### Limitations

This study has several limitations. It was not a randomised trial, with treatment allocation determined by clinical triage. Residual confounding by severity remains possible despite matching and adjustment: all complications and comorbidities were concentrated in DTC patients, indicating a degree of residual severity imbalance that recorded variables could not fully capture. Clinical complications occurring during treatment may also represent intermediate outcomes on the causal pathway from severity to death rather than independent confounders, meaning adjustment for them in the primary model may over-correct; the sensitivity analysis excluding complicated DTC patients should be interpreted with this in mind. The E-value of 4·40 quantifies the unmeasured confounding required to explain the treatment modality association, and the consistency of results across analytical approaches supports non-inferiority of HBC. Case ascertainment relied on clinical diagnosis. Laboratory confirmation was obtained for only a small minority of cases, with most classified as suspected diphtheria, which may have introduced diagnostic misclassification. The retrospective design introduces recall bias, though triangulation with clinical records mitigated this for key variables. Only 68·5% of sampled patients were traced, and selection bias is possible if those lost to follow-up differed systematically by treatment group or outcome. Because the cohort comprised only patients with mild disease, findings cannot be extrapolated to moderate or severe diphtheria, for which facility-based care with DAT remains essential. Finally, the safety of HBC cannot be assumed in settings without structured triage, follow-up, and referral systems, or for other pathogens.

### Implications for policy and response

These results suggest that HBC could be considered in epidemic response, provided triage reliably distinguishes mild from moderate or severe disease, antibiotic treatment is initiated upon presentation, and structured follow-up with a functioning referral pathway is in place. Under these conditions, HBC can prioritise hospital capacity and DAT supplies for patients at most need, reduce treatment delays, and achieve outcomes not detectably different from facility-based care.

Diphtheria continues to spread across West and Central Africa, with eight countries reporting active outbreaks as of late 2025.^17,18^ Global DAT supplies remain unstable, and threats to vaccination funding risk further erosion of coverage.^19^ In this context, evidence supporting safe and effective home-based management has implications beyond Kano State alone.

## Data Availability

Anonymised individual–level data will be made available upon reasonable request to the corresponding author, subject to approval by the MSF data sharing committee.

## Conclusions

In the context of a large diphtheria epidemic that overwhelmed hospital capacity in Kano State, Nigeria, home-based care for patients with mild disease produced clinical outcomes, household transmission levels, and patient acceptability comparable to facility-based treatment across multiple analytical approaches. The determinants of survival were the timing of treatment and vaccination status, not the setting of care. These findings support inclusion of decentralised care in diphtheria response frameworks, particularly where hospital capacity is limited.

## Acknowledgements

We thank the participating communities and local leaders of Dawakin Tofa, Fagge, Nassarawa, and Ungogo for their cooperation. We are grateful to the field investigators and data teams for their dedication, and to the MSF WaCA coordination for operational support. The study was conducted by Epicentre in collaboration with MSF WaCA and the Kano State Ministry of Health.

## Contributors

JP, EG and FA conceptualised the study and designed the methodology. JP, SH, KU, and FA led field planning and oversaw data collection. YK, PEE, HJOO, AKS, IIA, and COD contributed to study design and protocol development, and to data collection and field coordination. JP conducted the formal analysis and wrote the original draft. All authors contributed to critical review and revision of the manuscript for important intellectual content and approved the final version for submission. JP accepts overall responsibility for the work.

## Data sharing

Anonymised individual-level data will be made available upon reasonable request to the corresponding author, subject to approval by the MSF data sharing committee.

